# Changes in Age-Specific Pregnancy Prevalence Proportions over the Covid-19 Pandemic in Manicaland, Zimbabwe

**DOI:** 10.1101/2025.09.23.25336434

**Authors:** Simon Gregson, Louisa Moorhouse, Rufurwokuda Maswera, Sophie Bagnay, Blessing Tsenesa, Phyllis Mandizvidza, Rebekah Morris, Constance Nyamukapa

**Affiliations:** Imperial College London, Biomedical Research and Training Institute, Harare; Manicaland Centre for Public Health Research, Biomedical Research and Training Institute, Harare

**Keywords:** Fertility, Pregnancy Prevalence, Covid-19, Sexual Debut, Contraception, Zimbabwe

## Abstract

Past infectious disease outbreaks have altered population fertility in ways that contribute to changes in population age-structure and subsequent needs for public services. However, little is known about how fertility changed over the Covid-19 pandemic in African populations where birth registration systems typically remain weak. Using serial cross-sectional data from population surveys conducted in Manicaland, Zimbabwe, before, during, and towards the end of Covid-19 (2018-23), we found that women’s overall odds of current pregnancy had changed little at the height of Covid-19 but dropped following the relaxation of lockdown measures. In the middle of the pandemic, reductions in pregnancy rates in adolescents, associated with delays in sexual activity and marriage, offset increases in 25-34-year-old women linked to reduced use of contraception. Late in the pandemic, the overall odds of current pregnancy fell, with only a partial recovery in teenage sexual activity but a strong rebound in use of contraception.

## Introduction

Understanding the demographic impact of Covid-19 is critical to inform the development of plans to control future infectious disease outbreaks. Past and ongoing pandemics have had substantial impacts on fertility in affected populations. For example, between 1918 and 1919, the influenza pandemic caused a 13% drop in births rates in the USA (Chandra et al. 2018); in generalised HIV epidemics in sub-Saharan Africa, a 10% prevalence of HIV infection was estimated to reduce total fertility by approximately 4% (Zaba and Gregson 1998); and the 2014-2015 Ebola outbreak in Guinea resulted in a short-term reduction in use of family planning methods (Camara et al. 2017). Such changes are significant as they can affect the rate and pattern of population ageing, future health challenges, and economic growth (Aassve et al. 2020).

In the light of this experience with previous infectious disease outbreaks, early in the Covid-19 pandemic, it was recognised that SARS-Cov-2 infection, Covid-19 morbidity and mortality, and associated public health control measures had the potential to alter fertility (Aassve et al. 2020). Changes in proximate determinants of fertility including increased cohabitation and reduced access to effective family planning methods, due to Government mobility restrictions and fear of infection, could tend to increase birth rates in the short-term. Equally, other changes such as postponement of marriage and childbearing intentions, due to lockdown measures and economic pressures, and reduced fecundity in infected individuals could place downward pressures on population fertility. On balance, Aassve and colleagues hypothesised that, depending on their stages in the demographic transition, in urban areas in low and middle income countries (LMICs), upward pressures on fertility from reduced access to contraception were likely to be offset by downward pressures from economic losses and uncertainty; whilst, in rural areas, the upward pressures would be reinforced by disruptions to development resulting in increases in fertility rates (Aassve et al. 2020). In LMICs, overall, they concluded that the net effect of Covid-19 might be to increase fertility in the short-term.

In a subsequent study of monthly birth data from 38 higher-income countries (HICs) in the Human Fertility Database, covering the period up to September 2022, Sobotka and colleagues found evidence for distinct short-term swings in fertility associated with different phases of the Covid-19 pandemic (Sobotka et al. 2024). Bujard and Andersson recorded reductions in fertility in Sweden and Germany early in 2022 (Bujard and Andersson 2024). In further analyses by birth order, Burkimsher found that, in many European countries, fertility rates declined between 2019 and 2020, recovered in 2021, and declined again in 2022 (Burkimsher 2025). Upturns in higher birth orders drove the increases in 2021 – whilst flexible working, financial support and easing the work-life balance appeared to have motivated existing families to have another child, uncertainty and difficulty of meeting or being able to live with a partner inhibited those wanting to start a family (Burkimsher 2025). The changes in birth rates over Covid-19 in HICs have also been associated with elements of economic uncertainty including inflation, intervention stringency, and vaccination progress, but not unemployment (Winkler-Dworak, Zeman, and Sobotka 2024).

In an analysis of monthly birth registration data from 18 predominantly low- and middle-income countries (LMICs), no impact was observed in 4 countries, a short-term decline followed by a rebound was found in 6 countries, a longer decline was found in 6 countries, and a short temporary increase was recorded in 2 countries (Kim et al. 2024). No sub-Saharan African countries were included in the LMICs study due to limitations in birth registration data. One of the very few studies that have been reported in the region analysed longitudinal survey data from Burkina Faso, Kenya, Democratic Republic of the Congo (Kinshasa) [DRC], and Nigeria (Lagos) that covered the pre- and early-Covid-19 pandemic periods (Moreau et al. 2023). The study reported no changes in women’s exposure to unintended pregnancy risk, together with 5-9 per cent increases in contraceptive prevalence in Burkina Faso, Kenya and Lagos. A separate study in Kinshasa, analysed data on numbers of births in health facilities and recorded a 45 per cent increase in births seven months after the national Covid-19 lockdown was implemented (Feng et al. 2022). However, the extent to which population movements at the beginning of lockdown contributed to this increase was not made clear.

In this paper, we contribute to filling the gap in knowledge about changes in fertility in sub-Saharan African populations, that resulted from the Covid-19 pandemic, by addressing the following two objectives using serial cross-sectional data from three rounds of a representative open-cohort general population survey carried out in Zimbabwe’s eastern province of Manicaland:

1. To measure changes in total and age-specific pregnancy prevalence proportions over the course of the Covid-19 epidemic in a sub-Saharan African population; and
2. To investigate possible pathways through which Covid-19 and local responses to the pandemic affected age-specific pregnancy prevalence proportions differentially by measuring changes in the proximate determinants of fertility

## Data and methods

### Covid-19 in Zimbabwe

A national lockdown was implemented in Zimbabwe on March 30^th^, 2020 after detection of the first Covid-19 case in the country on March 17^th^. Four large waves of new infections followed with peaks in August 2020, January 2021, July 2021 and December 2021 (worldometer 2024). Relaxations and re-tightenings of the national lockdown (alternating between levels two and four) were made between these waves culminating in a steady easing of measures from early 2022 onwards (Morris et al. 2024). Lockdown measures included restrictions on movements, school closures, and bans on public gatherings including sports fixtures, church services and weddings (Murewanhema et al. 2022). The national vaccination programme was rolled out from February 2021 (Murewanhema et al. 2022; Morris et al. 2024). It is estimated that cumulative totals of 266,359 SARS-CoV-2 infections and 5,740 Covid-19 deaths occurred up to April 2024 in Zimbabwe (worldometer 2024) in a population estimated at 15.2 million people in the 2022 national census (Zimbabwe National Statistics Agency 2023).

### Study population and data collection

The data for the study were extracted from the three most recent consecutive rounds of a longitudinal general population open-cohort survey conducted in eight sub-communities in the Mutasa, Makoni, Nyanga and Mutare Urban districts in Manicaland province, Zimbabwe. The eight communities comprised two ‘high-density’ urban suburbs, two small towns, two agricultural estates, and two rural subsistence farming areas.

In each survey round, a prospective household census was updated and eligible individuals were identified. In the first (‘pre-’ Covid-19) survey, data were collected between July 2018 and December 2019 in face-to-face interviews; in the second (‘mid-’ Covid-19) survey, data were collected between February 2021 and July 2021 in telephone interviews; and in the third (‘late-’ Covid-19) survey, data were collected between July 2022 and March 2023 in face-to-face interviews (household census) and telephone interviews (individual survey). In last two surveys, eligibility for individual interviews was limited to household members aged 15 years and above usually resident in a random sample of two-thirds of the households enumerated in the corresponding census. In the pre-Covid-19 survey, young women (aged 15-24 years) and young men (15-29 years) were over-sampled as trials of interventions to increase use of pre-exposure prophylaxis (PrEP) and voluntary male medical circumcision (VMMC) were running in these groups. Therefore, for the pre-Covid-19 survey, all younger people were eligible to participate. Detailed information on the data collection procedures is available in previous publications (Gregson et al. 2017; Rao et al. 2022; Morris et al. 2024; Bagnay et al. Submitted) from the Manicaland Centre for Public Health Research (http://www.manicalandhivproject.org/).

In each survey, participants were requested to provide data on their socio-demographic characteristics, sexual behaviour, reproductive history, use of family planning methods and HIV infection and antiretroviral treatment (ART) status. In the first survey, provider-initiated testing and counselling for HIV infection (PITC) was included with consent for collection of dried blood spot samples for HIV testing being sought from those who declined PITC (e.g. due largely to their having previously received a positive HIV test result but being willing to participate in the study). In the last two surveys, additional data were collected on knowledge about Covid-19 and compliance with Government measures to control the spread of SARS-CoV-2 infection.

99.5 per cent (7568/7607), 87.6 per cent (6106/6970), and 97.3 per cent (7596/7808) of households identified in the survey areas were enumerated in the pre-, mid- and late-Covid-19 censuses respectively. 85.4 per cent (4652/5454), 89.4 per cent (3826/4281), and 86.2 per cent (4160/4828) of the women enumerated in these households in pre-, mid- and late-Covid-19 censuses and eligible for the current analyses (i.e. aged 15-49 years) chose to participate in the corresponding surveys.

The socio-demographic composition of the study population changed over the three surveys due to a combination of: 1) Covid-19-related migration; 2) changes in participation bias due to the changes in interview methods used in the surveys; and 3) the relative over-sampling of younger women in the pre-Covid-19 survey. After adjusting for the last of these three factors, the study population was slightly older in the mid- and late-Covid-19 surveys than in the pre-Covid-19 survey and was more concentrated in households in the second and third poorest wealth quintiles (Table 1). In the late-Covid-19 survey, the study population was more concentrated in urban communities than before Covid-19 (25.1 per cent *versus* 19.2 per cent; age-adjusted-odds ratio = 1.48, 95 per cent confidence interval (CI) 1.35-1.61) with a smaller proportion of women being in a marital union. HIV prevalence in the study population (women aged 15-49 years) in the pre-Covid-19 survey was 12.3 per cent (95 per cent CI 11.4–13.2) (Rao et al. 2022).

**Table 1.** Changes across survey rounds in the proportions of women aged 15-49 years in the population sample by socio-demographic characteristics, Manicaland, east Zimbabwe.

|  | Late-Covid-19 vs. Pre-Covid-19 |  |  |  | Mid-Covid-19 vs. Pre-Covid-19 |  |  |  | Pre-Covid-19 |  |
| --- | --- | --- | --- | --- | --- | --- | --- | --- | --- | --- |
|  | aOR<sup>1</sup> | 95 per cent CI | % | N | aOR<sup>1</sup> | 95 per cent CI | % | N | % | N |
| *Age-group* |  |  |  |  |  |  |  |  |  |  |
| 15-19 | 0.96 | 0.87-1.05 | 19.1 | 795 | 0.99 | 0.91-1.08 | 19.7 | 753 | 25.1 | 1167 |
| 20-24 | 0.92 | 0.83-1.02 | 17.2 | 714 | 0.83 | 0.75-0.92 | 15.4 | 588 | 22.5 | 1049 |
| 25-29 | 0.91 | 0.81-1.03 | 13.1 | 545 | 1.01 | 0.91-1.12 | 14.3 | 547 | 11.9 | 554 |
| 30-34 | 0.88 | 0.78-0.99 | 13.5 | 560 | 0.85 | 0.77-0.95 | 13.3 | 507 | 12.8 | 594 |
| 35-39 | 1.13 | 1.00-1.28 | 14.2 | 592 | 1.17 | 1.05-1.30 | 14.9 | 569 | 10.9 | 508 |
| 40-44 | 1.12 | 0.98-1.28 | 12.1 | 505 | 1.06 | 0.95-1.19 | 11.6 | 445 | 9.3 | 432 |
| 45-49 | 1.26 | 1.10-1.44 | 10.8 | 449 | 1.26 | 1.12-1.42 | 10.9 | 417 | 7.5 | 348 |
| *Location* |  |  |  |  |  |  |  |  |  |  |
| Urban | 1.48 | 1.35-1.61 | 25.1 | 1045 | 1.02 | 0.94-1.11 | 18.7 | 717 | 19.2 | 893 |
| Periurban | 0.89 | 0.82-0.96 | 28.4 | 1183 | 1.06 | 0.99-1.12 | 32.1 | 1230 | 30.6 | 1425 |
| Agricultural estate | 1.04 | 0.96-1.13 | 22.1 | 920 | 1.00 | 0.93-1.07 | 21.4 | 817 | 21.2 | 986 |
| Rural | 0.78 | 0.72-0.84 | 24.3 | 1012 | 0.93 | 0.87-0.99 | 27.8 | 1062 | 29.0 | 1348 |
| *Marital status* |  |  |  |  |  |  |  |  |  |  |
| Single | 1.21 | 1.06-1.38 | 23.2 | 966 | 1.15 | 1.01-1.31 | 22.9 | 875 | 27.2 | 1,265 |
| Married | 0.86 | 0.79-0.94 | 61.6 | 2,562 | 0.90 | 0.83-0.98 | 62.4 | 2,388 | 60.3 | 2,803 |
| Divorced or separated | 1.18 | 1.04-1.35 | 11.6 | 484 | 1.10 | 0.97-1.25 | 10.8 | 415 | 9.3 | 431 |
| Widowed | 0.81 | 0.66-1.01 | 3.6 | 148 | 0.88 | 0.73-1.06 | 3.9 | 148 | 3.3 | 153 |
| *Education<sup>2</sup>* |  |  |  |  |  |  |  |  |  |  |
| None/primary | 0.80 | 0.72-0.88 | 13.8 | 573 | 1.71 | 1.56-1.87 | 25.2 | 963 | 15.7 | 730 |
| Secondary/higher | 1.25 | 1.13-1.38 | 86.2 | 3587 | 0.59 | 0.53-0.64 | 74.8 | 2863 | 84.3 | 3922 |
| *Socio-economic status (quintile)* |  |  |  |  |  |  |  |  |  |  |
| Poorest | 0.59 | 0.49-0.69 | 4.9 | 202 | 0.76 | 0.65-0.88 | 6.8 | 262 | 8.8 | 411 |
| Second poorest | 1.14 | 1.05-1.25 | 40.3 | 1,678 | 1.15 | 1.05-1.25 | 42.4 | 1,624 | 40.1 | 1,864 |
| Third poorest | 1.16 | 1.05-1.27 | 28.1 | 1,170 | 1.24 | 1.13-1.37 | 28.2 | 1,080 | 24.0 | 1,118 |
| Fourth poorest | 0.88 | 0.80-0.97 | 25.3 | 1,051 | 0.76 | 0.69-0.84 | 21.6 | 826 | 25.5 | 1,185 |
| Least poor | 0.87 | 0.61-1.22 | 1.4 | 59 | 0.54 | 0.36-0.81 | 0.9 | 34 | 1.6 | 74 |
| *Church denomination* |  |  |  |  |  |  |  |  |  |  |
| Protestant | 0.97 | 0.89-1.06 | 21.0 | 872 | 0.91 | 0.83-0.99 | 20.1 | 769 | 21.8 | 1,015 |
| Roman Catholic | 0.90 | 0.78-1.04 | 6.7 | 277 | 1.03 | 0.91-1.16 | 7.9 | 302 | 7.6 | 353 |
| Pentecostal | 0.96 | 0.88-1.05 | 24.7 | 1,029 | 0.99 | 0.90-1.08 | 24.4 | 933 | 24.8 | 1,153 |
| Apostolic | 1.57 | 1.44-1.71 | 33.1 | 1,377 | 1.11 | 1.02-1.21 | 26.6 | 1,016 | 24.5 | 1,140 |
| Zionist | 0.85 | 0.73-0.98 | 6.2 | 256 | 0.82 | 0.71-0.93 | 5.8 | 222 | 7.0 | 324 |
| Other | 0.48 | 0.41-0.55 | 6.2 | 258 | 1.13 | 1.00-1.27 | 13.2 | 506 | 11.8 | 551 |
| None | 0.86 | 0.66-1.12 | 2.2 | 91 | 0.79 | 0.61-1.04 | 2.0 | 78 | 2.5 | 116 |
| *HIV infection status* |  |  |  |  |  |  |  |  |  |  |
| Infected |  | - |  |  |  | - |  |  | 10.9 | 483 |
| Uninfected |  | - |  |  |  | - |  |  | 89.1 | 3954 |
| *Birth history* |  |  |  |  |  |  |  |  |  |  |
| Nulli-parous | 1.24 | 1.08-1.42 | 25.1 | 1,046 | 1.29 | 1.13-1.47 | 25.3 | 969 | 29.8 | 1,384 |
| Parous | 0.81 | 0.71-0.93 | 74.9 | 3,114 | 0.77 | 0.68-0.88 | 74.7 | 2,857 | 70.2 | 3,268 |

<sup>1</sup> aOR = odds ratio adjusted for the effects of community location and age-group including over-sampling of women aged 15-24 years in the pre-Covid-19 survey.
<sup>2</sup> In the mid-Covid-19 survey, there was an error in the data capture system that led to some study participants with secondary education being misclassified as having received only primary education.

### Data analysis

Age-specific fertility rates (ASFRs) and total fertility rates (TFRs) were calculated using data collected on live births in the last 12 months in each of the three surveys. Age-specific pregnancy prevalence proportions (ASPPPs) per thousand women were calculated using self-reported data collected on current pregnancy at the time of the survey. An age-standardised pregnancy prevalence proportion (PPP) was also calculated. ASFRs and ASPPPs were calculated for 5-year age-groups and also for summarised age-groups (15-19 years, 20-24 years, 25-34 years, and 35-49 years). The TFRs and the age-standardised PPP were calculated for ages 15-49 years using the ASFRs and ASPPPs calculated for the 5-year age-groups. Odds ratios (aORs) with cluster-robust 95 per cent CIs for current pregnancy and recent birth in the mid-Covid-19 and late-Covid-19 surveys compared to the pre-Covid-19 survey, adjusted for the confounding effects of changes in age and community location between surveys, were estimated using multi-variable logistic regression models. This was done for all women in the study population and for socio-demographic sub-groups.

To examine the possible contributions of changes in the proximate determinants of fertility (Davis and Blake 1956; Bongaarts 1978) to changes in age-specific fertility observed over the course of the Covid-19 pandemic, figures were constructed, for each of the broader age-ranges of women, comparing changes between survey rounds in the proportions reporting: (1) ever having had sex, (2) currently being in a marital relationship; (3) currently abstaining from sexual intercourse (in those who had started sex); (4) consistently using a family planning method (in those who had started sex); (5) having had a birth in the last 12 months; and (6) currently being pregnant. 95 per cent CIs were calculated and displayed for each of these proportions to aid interpretation. Women reporting being married or in a long-term (≥12 months) or cohabiting relationship were taken to be in a marital relationship. Sexual abstinence was defined as not having had sex for 3 months or more. Consistent use of family planning methods was defined as using a modern method of contraception (pills, injectables, condoms, IUDs, implants and sterilisation) most or all of the time. For sexual abstinence and use of family planning methods in sexually experienced women, multi-variable logistic regression models, adjusted for population changes in age and community location between surveys, were constructed to estimate aORs for differences in these proximate determinants in the mid-Covid-19 and late-Covid-19 surveys compared to the pre-Covid-19 survey. Changes between Covid-19 periods in these proximate determinants were investigated in this way by marital status for sexually experienced women and by age-group and community location for currently married women. Multi-variable logistic regression models were also used to estimate aORs and cluster-robust 95 per cent CIs for use of different family planning methods in the mid-Covid-19 and late-Covid-19 surveys compared to the pre-Covid-19 survey.

All analyses were done using Stata version 17 (StataCorp. 2021).

### Ethical approval

Ethical approval for the Manicaland Study was provided by the Imperial College London Research Ethics Committee (20IC6436) and the Medical Research Council of Zimbabwe (MRCZ/A/2703).

## Results

### Changes in overall fertility rates over time in Manicaland, Zimbabwe

The TFR in the study communities in the pre-Covid-19 survey was 3.67 live births per women and the proportion of women with a birth in the year preceding this survey was 11.3 per cent (*n* = 4,652) (Table S1). The proportion of women pregnant at interview in the pre-Covid-19 survey was 6.2 per cent (*n* = 4,652) (Table 2), and was highest in women aged 20-24 years (9.4 per cent, *n* = 1,049), in those living on agricultural estates (8.3 per cent, *n* = 986) and in the poorest quintile of households (7.7 per cent, *n* = 411), in married women (8.6 per cent, *n* = 2,803), and in members of Apostolic (7.5 per cent, *n* = 1,140) and Zionist (9.7 per cent, *n* = 324) churches. Low proportions were found in single (1.2 per cent, *n* = 1,265) and widowed (2.0 per cent, *n* = 153) women. In the pre-Covid-19 survey, current pregnancy was less common overall in women living with HIV (WLHIV) (infected women on ART: 4.4 per cent, *n* = 313; women not on ART: 3.4 per cent, *n* = 170) than in uninfected women (6.5 per cent, *n* = 3,954) (Table 2). Pregnancy prevalence proportions in WLHIV were higher than for uninfected women at ages 20-24 years but lower than for uninfected women at ages 25-34 years (Figure 1C).

**Table 2.** Adjusted odds ratios for current pregnancy in late- and mid-Covid-19 compared to before Covid-19, by socio-demographic status, women aged 15-49 years, Manicaland, Zimbabwe.

| Characteristic | Late-Covid-19 vs. Pre-Covid-19 |  |  |  | Mid-Covid-19 vs. Pre-Covid-19 |  |  |  | Pre-Covid-19 |  |
| --- | --- | --- | --- | --- | --- | --- | --- | --- | --- | --- |
|  | AOR | 95% *per cent CI* | % | N | AOR | 95% *per cent CI* | % | N | % | N |
| *All women* | 0.73 | 0.60-0.88 | 4.4 | 4,160 | 0.97 | 0.81-1.17 | 5.8 | 3,826 | 6.2 | 4,652 |
| *Age-group* |  |  |  |  |  |  |  |  |  |  |
| 15-19 | 0.63 | 0.42-0.96 | 4.4 | 795 | 0.54 | 0.34-0.84 | 3.7 | 753 | 6.8 | 1,167 |
| 20-24 | 0.83 | 0.59-1.16 | 8.0 | 714 | 0.98 | 0.69-1.39 | 9.4 | 588 | 9.4 | 1,049 |
| 25-34 | 0.84 | 0.61-1.16 | 6.6 | 1,105 | 1.35 | 1.00-1.81 | 10.4 | 1,054 | 7.9 | 1,148 |
| 35-49 | 0.46 | 0.26-0.83 | 1.2 | 1,546 | 0.79 | 0.48-1.32 | 2.0 | 1,431 | 2.6 | 1,288 |
| *Location* |  |  |  |  |  |  |  |  |  |  |
| Urban | 0.84 | 0.53-1.31 | 3.8 | 1,045 | 1.25 | 0.79-1.96 | 5.4 | 717 | 4.6 | 893 |
| Periurban | 0.84 | 0.59-1.20 | 4.7 | 1,183 | 0.87 | 0.62-1.23 | 4.7 | 1,230 | 5.7 | 1,425 |
| Agricultural estate | 0.59 | 0.40-0.86 | 4.9 | 920 | 1.06 | 0.76-1.49 | 8.3 | 817 | 8.3 | 986 |
| Rural | 0.70 | 0.48-1.02 | 4.3 | 1,012 | 0.86 | 0.61-1.22 | 5.4 | 1,062 | 6.3 | 1,348 |
| *Marital status* |  |  |  |  |  |  |  |  |  |  |
| Single | 0.35 | 0.11-1.14 | 0.4 | 966 | 0.87 | 0.36-2.11 | 0.9 | 875 | 1.2 | 1,265 |
| Married | 0.87 | 0.71-1.07 | 6.6 | 2,562 | 1.16 | 0.95-1.42 | 8.6 | 2,388 | 8.6 | 2,803 |
| Divorced or separated | 0.53 | 0.24-1.17 | 2.1 | 484 | 0.41 | 0.15-1.09 | 1.5 | 415 | 3.9 | 431 |
| Widowed | - | - | 0.0 | 148 | - | - | 1.4 | 148 | 2.0 | 153 |
| *Sexual activity* |  |  |  |  |  |  |  |  |  |  |
| Sexual debut | 0.79 | 0.65-0.95 | 5.4 | 3,381 | 1.08 | 0.89-1.30 | 7.2 | 3,102 | 7.5 | 3,679 |
| *Socio-economic status (quintile)* |  |  |  |  |  |  |  |  |  |  |
| Poorest | 0.77 | 0.37-1.59 | 6.4 | 202 | 0.96 | 0.51-1.80 | 6.9 | 262 | 7.7 | 411 |
| Second poorest | 0.73 | 0.55-0.97 | 4.8 | 1,678 | 1.04 | 0.80-1.36 | 6.5 | 1,624 | 6.7 | 1,864 |
| Third poorest | 0.67 | 0.46-0.97 | 4.4 | 1,170 | 0.90 | 0.63-1.27 | 5.7 | 1,080 | 6.4 | 1,118 |
| Fourth poorest | 0.67 | 0.43-1.04 | 3.2 | 1,051 | 0.85 | 0.55-1.32 | 4.1 | 826 | 4.9 | 1,185 |
| Least poor | 3.21 | 0.53-19.4 | 5.1 | 59 | 3.03 | 0.40-23.5 | 5.9 | 34 | 3.2 | 74 |
| *Church denomination* |  |  |  |  |  |  |  |  |  |  |
| Protestant | 1.01 | 0.64-1.60 | 4.1 | 872 | 1.09 | 0.69-1.73 | 4.4 | 769 | 4.2 | 1,015 |
| Roman Catholic | 0.66 | 0.27-1.63 | 2.9 | 277 | 1.42 | 0.67-3.01 | 5.6 | 302 | 4.3 | 353 |
| Pentecostal | 0.72 | 0.49-1.06 | 4.3 | 1,029 | 0.98 | 0.68-1.41 | 5.8 | 933 | 6.0 | 1,153 |
| Apostolic | 0.61 | 0.44-0.86 | 4.4 | 1,377 | 0.98 | 0.71-1.37 | 7.1 | 1,016 | 7.5 | 1,140 |
| Zionist | 0.56 | 0.28-1.11 | 5.5 | 256 | 0.64 | 0.32-1.27 | 5.9 | 222 | 9.7 | 324 |
| Other | 0.80 | 0.42-1.51 | 5.4 | 258 | 0.77 | 0.45-1.29 | 5.1 | 506 | 6.6 | 551 |
| None | 1.36 | 0.46-4.03 | 6.6 | 91 | 1.79 | 0.58-5.51 | 7.7 | 78 | 6.8 | 116 |
| *HIV infection status* |  |  |  |  |  |  |  |  |  |  |
| Infected - on ART |  |  | - |  |  |  | - |  | 4.4 | 313 |
| Infected - no ART |  |  | - |  |  |  | - |  | 3.4 | 170 |
| Uninfected |  |  | - |  |  |  | - |  | 6.5 | 3,954 |
| *Birth history* |  |  |  |  |  |  |  |  |  |  |
| Nulli-parous | 0.62 | 0.45-0.86 | 5.7 | 1,046 | 0.67 | 0.48-0.92 | 5.9 | 969 | 8.4 | 1,384 |
| Parous | 0.77 | 0.61-0.98 | 3.9 | 3,114 | 1.14 | 0.91-1.42 | 5.8 | 2,857 | 5.5 | 3,268 |

AOR: Odds ratios from multivariable logistic regression adjusted for age-group and community location.

**Figure 1.**
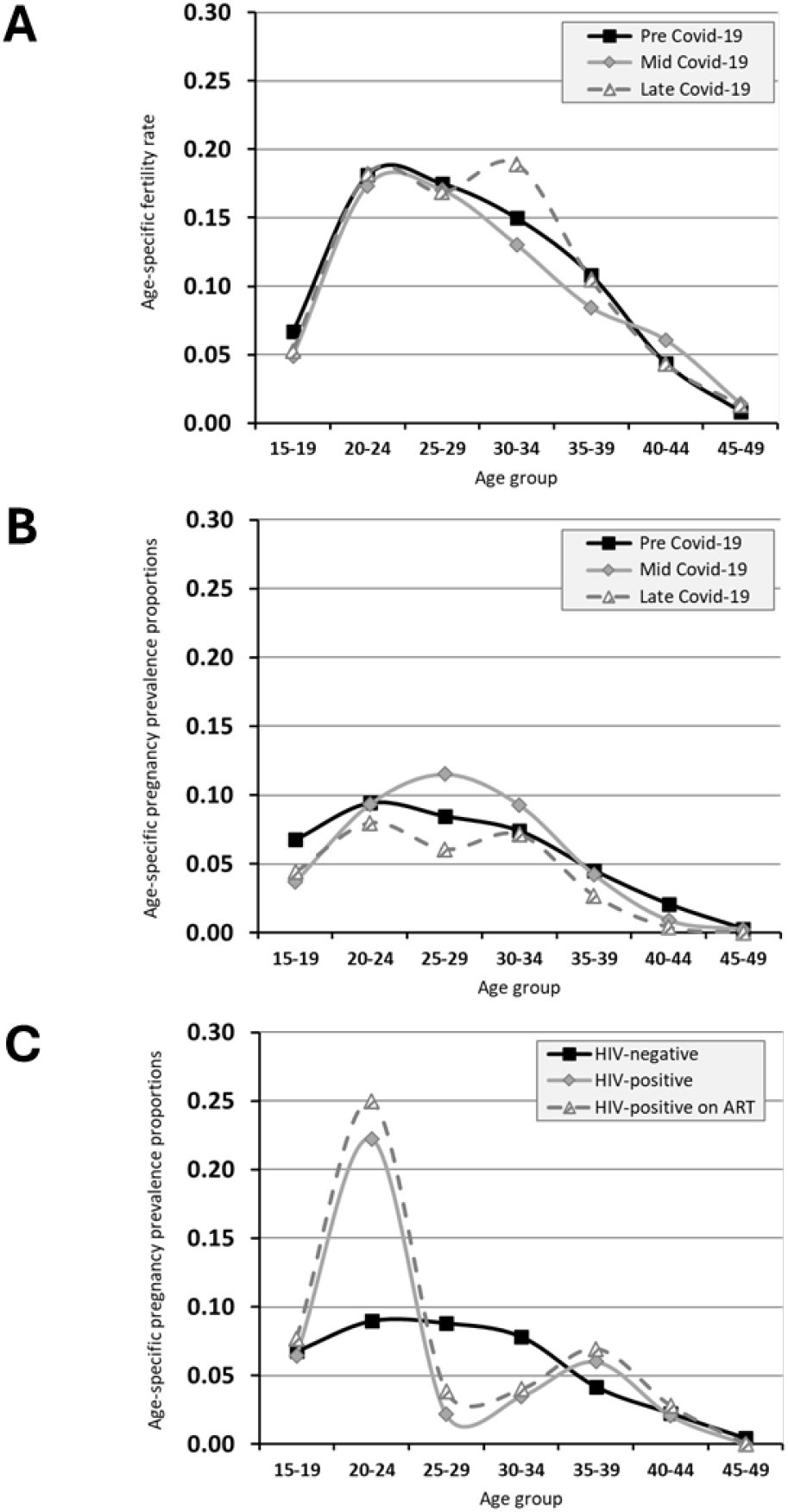
Age-specific fertility rates (graph A) and age-specific pregnancy rates (graph B) by stage of the Covid-19 pandemic and by HIV infection status prior to the (graph C), Manicaland, Zimbabwe.

The TFR fell to 3.41 in the mid-Covid-19 survey before recovering to 3.77 in the late-Covid-19 survey. However, the reduction in odds of a recent birth between the pre- and mid-Covid-19 surveys was not statistically significant (9.9 per cent *versus* 11.3 per cent; aOR = 0.90, 95 per cent CI, 0.78-1.04) (Table S1). The age-standardised pregnancy prevalence proportion also remained unchanged between the pre-Covid-19 (55.7 per thousand women) and mid-Covid-19 surveys (56.0 per thousand) but was lower in the late-Covid-19 survey (41.0 per thousand).

The reduced age-adjusted odds of current pregnancy in the late-Covid-19 period was most evident in the agricultural estates but also showed a trend in the rural communities (Table 2). After adjusting for age and community location, reduced age-adjusted odds of current pregnancy in the late-Covid-19 period were found in single women (0.4 per cent *versus* 1.2 per cent; aOR = 0.35, 95 per cent CI, 0.11-1.14), women living in the second and third poorest quintiles of households, women in Apostolic churches, and in nulli-parous women (Table 2). Women who had not previously had a live birth when interviewed in the mid-Covid-19 survey also had lower current pregnancy rates than their counterparts interviewed in the pre-Covid-19 survey.

### Age-specific pregnancy prevalence proportions

The study data on age-specific pregnancy prevalence proportions by Covid-19 period are shown in Table 2 and Figure 1B. Pregnancy proportions in 15-19 year-old women were substantially reduced in both the mid-Covid-19 period and the late-Covid-19 period. Non statistically significant reductions in recent births were also found in this age-group in the mid- and late-Covid-19 periods (Table S1). Pregnancy prevalence proportions in 20-24 year-old women did not change between the pre- and mid-Covid-19 survey periods but showed a non-significant reduction in the late-Covid-19 period. In the 25-34 year age-group, the pregnancy prevalence proportion increased in the middle of the Covid-19 outbreak but fell back to pre-Covid-19 levels in the late-Covid-19 period. In the 35-49 year age-group, the pregnancy prevalence proportion didn’t change in the midst of Covid-19 but was lower than before Covid-19 in the late-Covid-19 period.

### Age-specific changes in the proximate determinants of fertility

In the 15-19-year age-range, fewer young women had started sex and were currently married in the mid-Covid-19 period compared to the pre-Covid-19 period (Figure 2). In those who had started sex, sexual abstinence increased, and use of family planning methods declined. Therefore, an overall reduction in sexual activity, partly linked to delayed married, seems to have been the main contributor to the reductions in fertility in this age-group. These changes had all been partially reversed as Covid-19 control measures were eased later in the pandemic. In 20-24-year-old women, there were smaller reductions in sexual debut and ever-marriage, an increase in divorce and separation, and a small increase in sexual abstinence during Covid-19; but their combined effects on fertility in this age-group were cancelled out by a drop in use of family planning methods. In 25-34-year-old women, sexual abstinence rose but use of family methods fell in the mid-Covid-19 survey. In the late Covid-19 period, sexual abstinence rose further, divorce and separation increased, and use of family planning methods recovered in this age-group. Changes in the proximate determinants of fertility in 35-49-year-old women generally were small over the course of the Covid-19 pandemic. Current marriage increased slightly during Covid-19 and use of family planning methods rose in the late-Covid-19 period.

**Figure 2.**
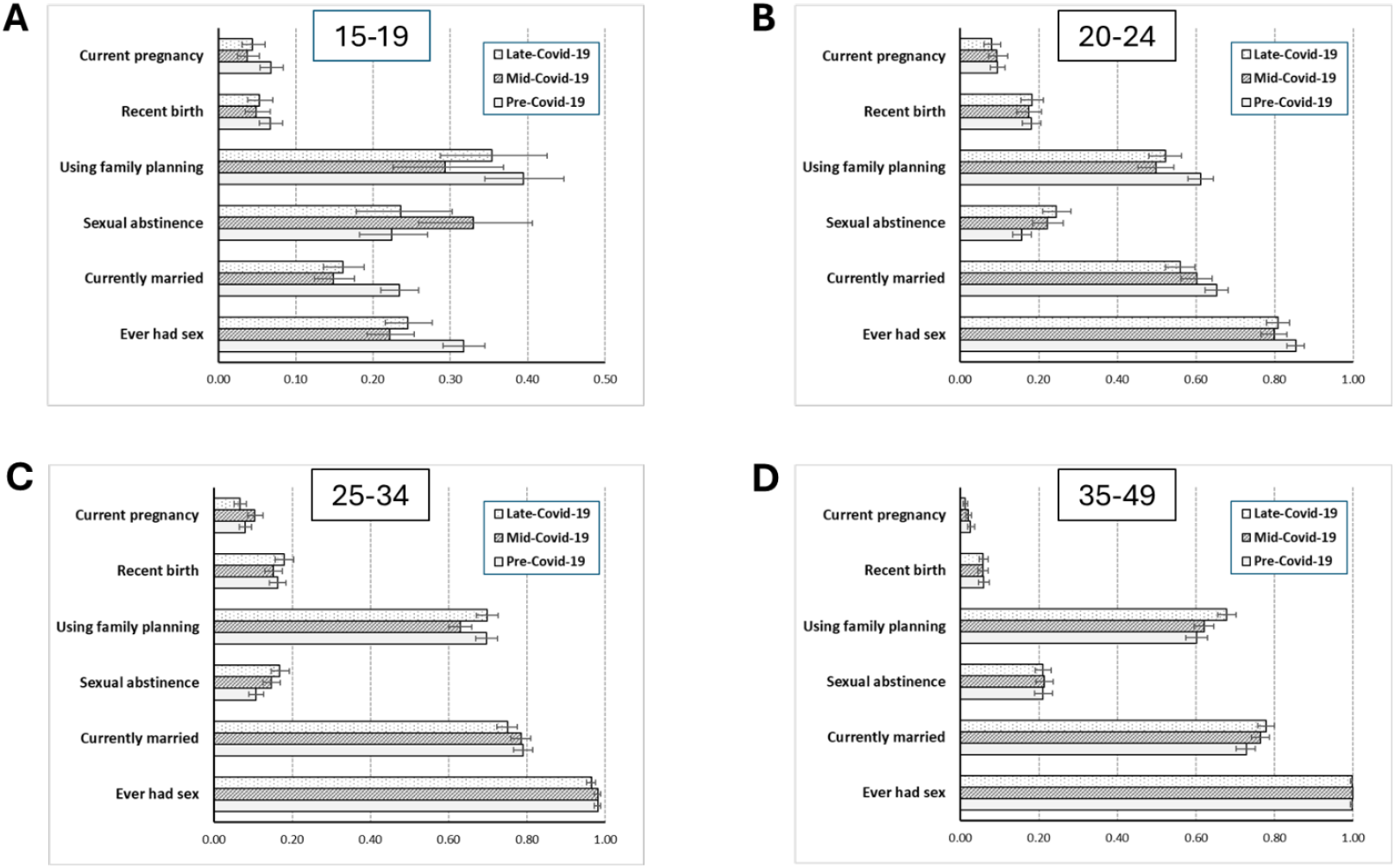
Changes over the course of the Covid-19 pandemic in proportions of women aged 15-49 years experiencing a birth in the last year and a current pregnancy, and in selected proximate determinants of fertility by age-group, Manicaland, Zimbabwe. Pre-Covid-19: July 2018 to December 2019; Mid-Covid-19: February to July 2021; Late-Covid-19: July 2022 to March 2023.

Overall, sexual abstinence amongst sexually-experienced women was more common in the mid-Covid-19 period (19.9 per cent; aOR = 1.27, 95 per cent CI, 1.14-1.42; *n* = 3,102) and in the late-Covid-19 period (20.4 per cent; aOR = 1.31, 95 per cent CI, 1.17-1.47; *n* = 3,381) than before Covid-19 (16.6 per cent, *n* = 3,679) (Table S2). This pattern was seen in all marital status groups except for widowed women for whom the sample sizes were smaller. In married women, the pattern was most evident in the peri-urban communities.

Consistent use of any family planning method was lower in the mid-Covid-19 period (58.8 per cent; aOR = 0.82, 95 per cent CI, 0.74-0.90; *n* = 3,102) than before Covid-19 (62.1 per cent, *n* = 3.679) (Table S2 and Table 3). However, this change was only statistically significant in currently married women, ages 20-34 years, and in those living on agricultural estates and in rural areas.

**Table 3.** Adjusted odds ratios for use of different types of family planning methods in late- and mid-Covid-19 compared to before Covid-19, sexually-experienced women aged 15-49 years, Manicaland, Zimbabwe.

|  | Late-Covid-19 vs. Pre-Covid-19 |  |  | Mid-Covid-19 vs. Pre-Covid-19 |  |  | Pre-Covid-19 |
| --- | --- | --- | --- | --- | --- | --- | --- |
|  | AOR | 95% *per cent CI* | % | AOR | 95% *per cent CI* | % | % |
| Main family planning methods used |  |  |  |  |  |  |  |
| Pills | 0.99 | 0.90-1.08 | 35.7 | 0.78 | 0.71-0.89 | 31.0 | 35.8 |
| Injectables | 0.97 | 0.87-1.09 | 17.9 | 0.70 | 0.62-0.80 | 13.9 | 18.5 |
| Male condoms | 0.95 | 0.81-1.12 | 8.3 | 0.90 | 0.76-1.07 | 7.8 | 8.3 |
| Female condoms | 0.66 | 0.36-1.18 | 0.35 | 1.04 | 0.62-1.75 | 0.90 | 0.79 |
| Sterilization | 2.18 | 1.19-3.98 | 1.1 | 3.44 | 1.94-6.08 | 1.6 | 0.46 |
| Other<sup>1</sup> | 1.89 | 1.47-2.43 | 5.0 | 2.75 | 2.17-3.49 | 7.2 | 2.6 |
| Any modern method<sup>2</sup> | 1.05 | 0.96-1.16 | 63.2 | 0.82 | 0.74-0.90 | 57.9 | 61.3 |
| Safe period | 0.13 | 0.02-1.00 | 0.03 | 0.14 | 0.02-1.06 | 0.03 | 0.23 |
| Withdrawal | 0.62 | 0.27-1.43 | 0.27 | 0.49 | 0.21-1.17 | 0.19 | 0.37 |
| Other | 0.96 | 0.50-1.84 | 0.55 | 1.28 | 0.70-2.35 | 0.74 | 0.56 |
| Any method<sup>2</sup> | 1.05 | 0.95-1.15 | 63.9 | 0.82 | 0.74-0.90 | 58.8 | 62.1 |
| N |  |  | 3,381 |  |  | 3,102 | 3,679 |

<sup>1</sup>Includes IUDs and implants.
<sup>2</sup>Some survey participants reported more than one main family planning method.
AOR: Odds ratios from multivariable logistic regression adjusted for age-group and community location.

### Changes in family planning methods over the course of the Covid-19 pandemic

Consistent use of modern methods of family planning by sexually-experienced women fell from 61.3 per cent in the Covid-19 survey to 57.9 per cent in the mid-Covid-19 survey (aOR = 0.82, 95 per cent CI, 0.74-0.90) before recovering to 63.2 per cent in the late-Covid-19 survey (aOR = 1.05, 95 per cent CI, 0.96-1.16) (Table 3). Contraceptive pills and injections were less commonly reported in the mid-Covid-19 survey than in the pre-Covid-19 survey. However, other modern methods (including IUDs and implants) and sterilisation were more frequently reported during Covid-19. All modern methods returned to pre-Covid-19 levels in the late Covid-19 survey except that use of sterilization and other methods remained higher after Covid-19 than before Covid-19.

## Discussion and conclusions

The drop in the TFR in women aged 15-49 years in Manicaland, east Zimbabwe, between the pre-Covid19 and mid-Covid-19 surveys – most pronounced in urban communities – suggests a possible short-lived reduction in new pregnancies very early in the pandemic. Thereafter, the overall pregnancy prevalence proportion remained stable at the height of the Covid-19 outbreak. However, below the surface, counteracting changes occurred in age-specific pregnancy prevalence proportions that could be accounted for by differential changes in proximate determinants of fertility. These changes in proximate determinants, in turn, likely resulted from spontaneous and Government-led responses to the Covid-19 pandemic. Teenage pregnancies dropped due to reductions in sexual activity and delays in marriage – probably due to fear of Covid-19 and Government restrictions on movements and social gatherings including weddings. However, the impact that this would have had on overall pregnancy rates was offset by an increase in pregnancies in 25-34 year-old women for whom the effect of a reduction in use of family planning methods – reflecting reduced availability and the Government restrictions on movement – seems to have outweighed the effect of a small increase in sexual abstinence.

Later in the local epidemic, as more infectious but less fatal strains of the Covid-19 virus evolved and took hold (Murewanhema and Mutsigiri-Murewanhema 2021), vaccination coverage increased (Murewanhema et al. 2022), and Covid-19 lockdown restrictions were eased (Morris et al. 2024), the overall pregnancy prevalence proportion declined. Teenage pregnancy rates remained low, reflecting continuing delays in marriage and onset of sexual activity – perhaps due to residual fear of Covid-19 morbidity and mortality and the accumulated economic impact of the epidemic – and levels of use of family planning methods recovered across all age-groups.

The data from Manicaland provide limited support for the *a priori* hypotheses suggested by Aassve and colleagues early in the pandemic (Aassve et al. 2020). Sample sizes are, of course, smaller when disaggregated into socio-economic settings but no statistically significant changes in pregnancy prevalence proportions were found in either urban or rural communities. There was little evidence for any overall change in pregnancy rates in the urban communities; but, in the rural communities, pregnancy rates became progressively lower from the pre-Covid-19 survey through to the late-Covid-19 survey.

In the one previous study reporting data on changes in pregnancy and birth rates in sub-Saharan African populations during Covid-19, Backhaus found declines in birth events in Burkina Faso, DR Congo, Kenya and Nigeria at the beginning of the outbreak (Backhaus 2022, 2024). As in Manicaland, in three of the four countries studied, teenage pregnancies declined early in Covid-19 (late 2020/early 2021) but no breakdown by age was reported for the most recent survey (late 2021/early 2022) and, as yet, no data have been published for later stages of the pandemic.

Beyond this, some other inter-related comparisons can be made. First, the TFR estimate in the late-Covid-19 survey (3.77 live births per woman) in Manicaland is quite close to the TFR recorded for Zimbabwe as a whole (3.9) in the Zimbabwe Demographic and Health Survey 2023/24 (ZDHS) (Zimbabwe National Statistics Agency and ICF 2025), for which the reference periods for recording births overlap, taking into account the shorter reference period used in the current study (i.e. 12 months *versus* 3 years). The TFR estimate for Manicaland province in the ZDHS is higher (4.6) however. This may reflect the more rural make-up of the province as a whole than is captured in the selectively more urban community locations covered by the current study.

A scoping review of 55 studies carried out in LMICs early in the Covid-19 pandemic found almost universal declines in contraceptive service provision (Polis et al. 2022). In Africa, Yu and colleagues recorded a marginal increase in contraceptive uptake very early in the Covid-19 pandemic in Kenya and Burkina Faso (Yu et al. 2024); but, more consistent with the current paper’s finding of a temporary reduction in use during Covid-19 in Manicaland, they also found evidence for a decrease in regions with high numbers of Covid-19 cases. In a qualitative study in Harare, Mashonaland East, and Bulawayo, young people (ages 16-24 years) were found to have disengaged from using contraceptives during Covid-19 following temporary closure of youth-friendly services (Mavodza et al. 2022). In Senegal, a similar change in the mix of family methods was found to that seen in Manicaland – i.e. away from use of shorter-acting methods (OCPs and injectables) towards use of long-acting methods (IUDs and implants) (Fuseini et al. 2022).

An interesting side finding in the pre-Covid-19 survey in Manicaland is the similarity in pregnancy rates between HIV-positive and HIV-negative teenagers. In earlier studies in the same study areas (Terceira et al. 2003) and elsewhere (Lewis et al. 2004), fertility rates generally have been higher in HIV-positive teenagers due to the selection for early unprotected sexual activity. A possible explanation for the disappearance of this differential could be an increase in survival to late adolescence amongst girls infected through mother-to-child transmission following the scale-up of paediatric ART services (Ferrand et al. 2010). Furthermore, the switch to initiating people living with HIV onto ART at early stages of infection in 2015 (World Health Organisation 2015) may have contributed to the apparent disappearance of the sub-fertility previously observed amongst older women living with HIV (Lewis et al. 2004; Yeatman et al. 2016).

Strengths of the current study include the availability and use of general population data collected from the same communities in four principal socio-economic strata in Zimbabwe’s Manicaland province before, during and towards the end of the Covid-19 pandemic. The survey questionnaires captured data that permitted calculation of short-term measures of fertility and of most of the proximate determinants of fertility that could plausibly have been affected by Covid-19 and local responses to the pandemic. An exception to this is that it has not been possible to investigate any effects of SARS-CoV-2 infection or vaccination on fertility. Reliable testing for SARS-Cov-2 infection was not widely available to the general public in Zimbabwe for much of the national epidemic and funding has not been available for specimen collection and testing to be done in the surveys. However, to date, there is no clear evidence for effects of Covid-19 or associated vaccines on either female or male fecundity (Wang et al. 2023; Javadzadeh et al. 2024; Gullo et al. 2024; Zace et al. 2022). Other important limitations include the use of self-reported data on current pregnancy status, and the change in interview method between surveys – from face-to-face interviews to telephone interviews – and the lower participation rate in the mid-Covid-19 census which may have introduced a change in participation bias. The latter was addressed partially by including controls in the analysis for community location and age-group.

Given that the lower overall pregnancy rates in the late-Covid-19 survey appeared to be due largely to lingering delays in onset of sexual activity at young ages, it seems possible that a modest baby boom could occur in the years following the pandemic as the economy recovers and women in the affected cohorts and in the cohorts that follow immediately after them marry and return to previous patterns of behaviour. Where possible, it will be helpful to carry out further surveys to establish the extent to which this is the case. The fast rebound in reported use of contraceptives suggests that the Zimbabwe National Family Planning Council was quick to restore services as lockdown measures were relaxed. However, in planning for possible future infectious disease outbreaks, it will be important to include measures that ensure that women – and especially teenage women – have the necessary knowledge and ability to continue to safely access contraception (Yu et al. 2024; Mavodza et al. 2022).

## Supporting information

Complete supplementary material document

## Data Availability

Data from the Manicaland Study can be obtained from the project website (http://www.manicalandhivproject.org/data-access.html). Here we provide a core data set which contains a sample of sociodemographic, sexual behaviour and HIV testing variables from all nine rounds of the main survey, as well as data used in the production of recent academic publications. If further data are required, a data request form must be completed (available to download from our website) and submitted to. If the proposal is approved, we will send a data sharing agreement which must be agreed upon before we release the requested data.

## Acknowledgements

We are very grateful to the Manicaland Centre for Public Health Research team and to the communities and study participants in Manicaland, east Zimbabwe for their kind assistance with the research.

