## Supplementary material for "Changes in Age-Specific Pregnancy Prevalence Proportions over the Covid-19 Pandemic in Manicaland, Zimbabwe": Complete supplementary material document

**Figure S1** Household census and individual survey participation rates in the Manicaland General Population Survey, 2018 to 2023. Graph A: pre-Covid-19, graph B: mid-Covid-19, and graph C: late-Covid-19.

**A**

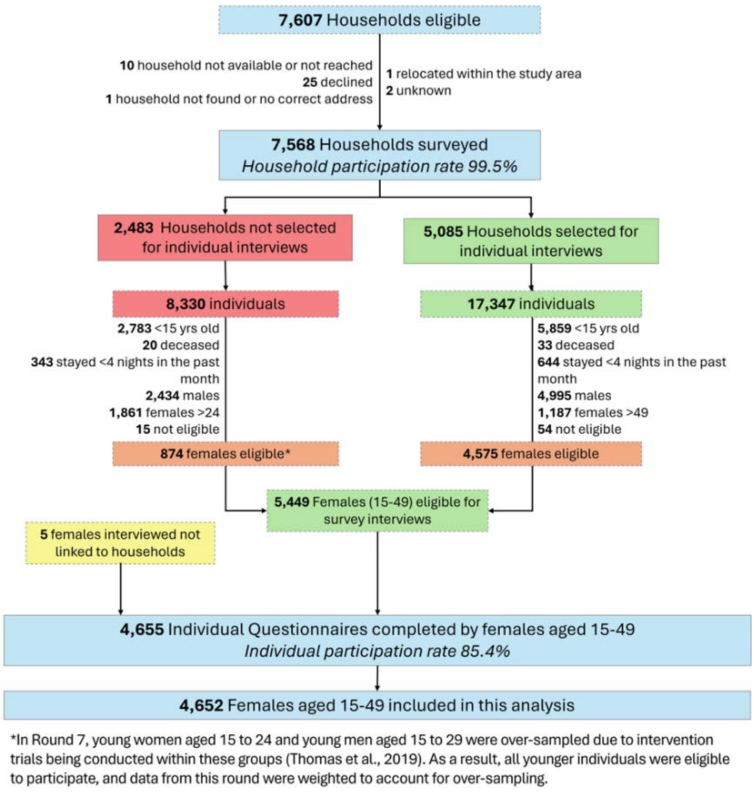

**B**

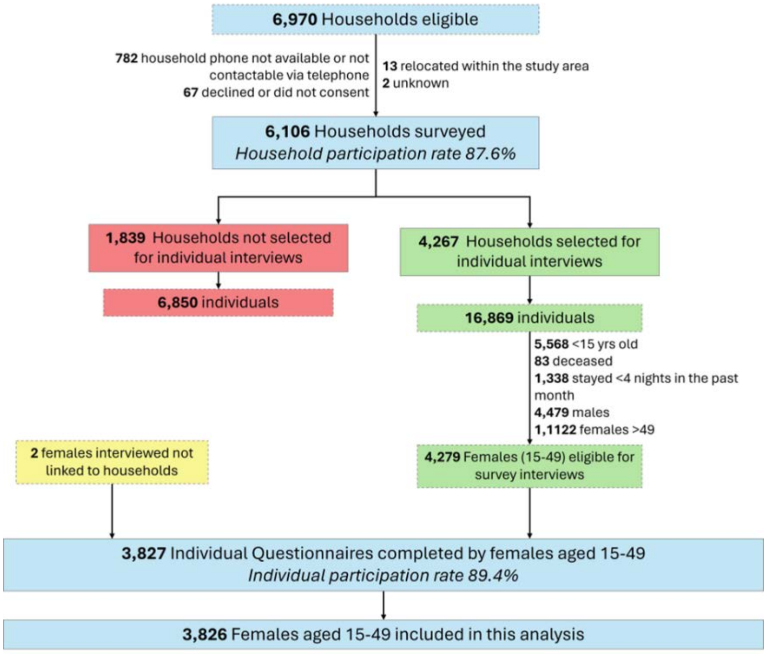
6,970 Households eligible
782 household phone not available or not contactable via telephone
67 declined or did not consent
13 relocated within the study area
2 unknown
6,106 Households surveyed
Household participation rate 87.6%
1,839 Households not selected for individual interviews
6,850 individuals
4,267 Households selected for individual interviews
16,869 individuals
5,568 <15 yrs old
83 deceased
1,338 stayed <4 nights in the past month
4,479 males
1,1122 females >49
2 females interviewed not linked to households
4,279 Females (15-49) eligible for survey interviews
3,827 Individual Questionnaires completed by females aged 15-49
Individual participation rate 89.4%
3,826 Females aged 15-49 included in this analysis

**C**

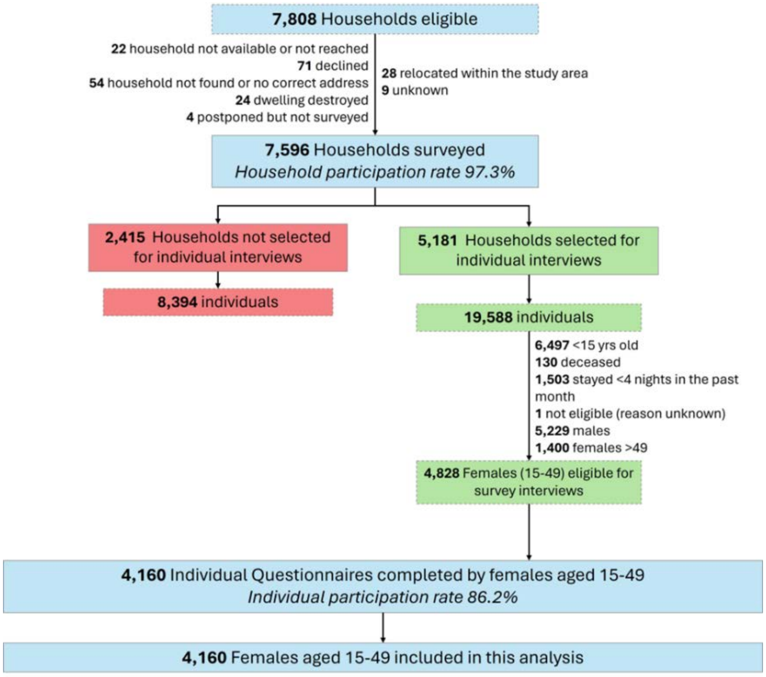
7,808 Households eligible
22 household not available or not reached
71 declined
54 household not found or no correct address
24 dwelling destroyed
4 postponed but not surveyed
28 relocated within the study area
9 unknown
7,596 Households surveyed
Household participation rate 97.3%
2,415 Households not selected for individual interviews
8,394 individuals
5,181 Households selected for individual interviews
19,588 individuals
6,497 <15 yrs old
130 deceased
1,503 stayed <4 nights in the past month
1 not eligible (reason unknown)
5,229 males
1,400 females >49
4,828 Females (15-49) eligible for survey interviews
4,160 Individual Questionnaires completed by females aged 15-49
Individual participation rate 86.2%
4,160 Females aged 15-49 included in this analysis

**Table S1** Adjusted odds ratios of a recent birth in late- and mid- Covid-19 compared to before Covid-19, women aged 15-49 years, Manicaland, Zimbabwe

| | Late-Covid-19 *vs.* Pre-Covid-19 | | | | Mid-Covid-19 *vs* . Pre-Covid-19 | | | | Pre-Covid-19 | |
|---|---|---|---|---|---|---|---|---|---|---|
| Characteristic | AOR | *95% per cent CI* | % | *N* | AOR | *95% per cent CI* | % | *N* | % | *N* |
| *All women* | 1.03 | 0.90-1.18 | 11.1 | 4,160 | 0.90 | 0.78-1.04 | 9.9 | 3,826 | 11.3 | 4652 |
| *Age-group* | | | | | | | | | | |
| 15-19 | 0.77 | 0.52-1.13 | 5.3 | 795 | 0.74 | 0.49-1.11 | 4.9 | 753 | 6.7 | 1,167 |
| 20-24 | 1.01 | 0.79-1.30 | 18.2 | 714 | 0.95 | 0.73-1.23 | 17.4 | 588 | 18.1 | 1,049 |
| 25-34 | 1.16 | 0.93-1.45 | 17.9 | 1,105 | 0.92 | 0.73-1.16 | 15.1 | 1,054 | 16.2 | 1,148 |
| 35-49 | 1.03 | 0.75-1.41 | 5.8 | 1,546 | 0.98 | 0.71-1.35 | 5.7 | 1,431 | 5.9 | 1,288 |
| *Location* | | | | | | | | | | |
| Urban | 0.84 | 0.63-1.12 | 10.6 | 1,045 | 0.63 | 0.45-0.88 | 8.0 | 717 | 12.7 | 893 |
| Periurban | 1.22 | 0.93-1.60 | 10.1 | 1,183 | 1.26 | 0.96-1.64 | 10.2 | 1,230 | 9.2 | 1,425 |
| Agricultural estate | 1.23 | 0.93-1.63 | 13.2 | 920 | 0.98 | 0.73-1.32 | 10.8 | 817 | 11.0 | 986 |
| Rural | 0.86 | 0.66-1.11 | 10.8 | 1,012 | 0.79 | 0.61-1.02 | 10.3 | 1,062 | 12.8 | 1,348 |
| *Marital status* | | | | | | | | | | |
| Single | 0.57 | 0.29-1.13 | 1.2 | 966 | 0.92 | 0.49-1.74 | 1.8 | 875 | 2.1 | 1,265 |
| Married | 1.13 | 0.97-1.31 | 15.3 | 2,562 | 0.96 | 0.81-1.12 | 13.5 | 2,388 | 15.4 | 2,803 |
| Divorced or separated | 1.38 | 0.87-2.17 | 11.2 | 484 | 1.00 | 0.61-1.65 | 8.4 | 415 | 8.8 | 431 |
| Widowed | 0.19 | 0.03-1.29 | 0.7 | 148 | 1.05 | 0.27-4.13 | 3.4 | 148 | 3.5 | 153 |
| *Sexual activity* | | | | | | | | | | |
| Sexual debut | 1.10 | 0.96-1.26 | 13.6 | 3,381 | 0.96 | 0.83-1.11 | 12.1 | 3,102 | 13.5 | 3679 |
| *Socio-economic status (quintile)* | | | | | | | | | | |
| Poorest | 1.15 | 0.59-1.65 | 11.4 | 202 | 0.99 | 0.66-1.98 | 10.3 | 262 | 12.1 | 411 |
| Second poorest | 1.03 | 0.84-1.27 | 12.2 | 1,678 | 0.81 | 0.66-1.01 | 10.0 | 1,624 | 12.5 | 1,864 |
| Third poorest | 1.05 | 0.80-1.38 | 11.8 | 1,170 | 1.11 | 0.85-1.45 | 11.9 | 1,080 | 11.0 | 1,118 |
| Fourth poorest | 0.91 | 0.68-1.22 | 8.4 | 1,051 | 0.79 | 0.57-1.10 | 7.3 | 826 | 9.5 | 1,185 |
| Least poor | 1.71 | 0.56-5.30 | 11.9 | 59 | 0.31 | 0.02-4.57 | 2.9 | 34 | 7.4 | 74 |
| *Church denomination* | | | | | | | | | | |
| Protestant | 1.10 | 0.80-1.51 | 9.5 | 872 | 1.12 | 0.80-1.57 | 9.4 | 769 | 8.8 | 1,015 |
| Roman Catholic | 0.64 | 0.36-1.13 | 7.9 | 277 | 0.59 | 0.34-1.01 | 7.3 | 302 | 11.5 | 353 |
| Pentecostal | 0.94 | 0.70-1.25 | 10.0 | 1,029 | 0.76 | 0.56-1.04 | 8.5 | 933 | 11.3 | 1,153 |
| Apostolic | 1.17 | 0.92-1.48 | 13.4 | 1,377 | 0.96 | 0.74-1.25 | 11.5 | 1,016 | 12.4 | 1,140 |
| Zionist | 0.78 | 0.45-1.35 | 9.4 | 256 | 0.94 | 0.54-1.64 | 10.8 | 222 | 12.1 | 324 |
| Other | 1.01 | 0.64-1.60 | 13.6 | 258 | 0.77 | 0.51-1.15 | 10.7 | 506 | 12.7 | 551 |
| None | 0.83 | 0.32-2.16 | 9.9 | 91 | 1.51 | 0.62-3.71 | 14.1 | 78 | 11.5 | 116 |
| *HIV infection status* | | | | | | | | | | |
| Infected - on ART | | - | | | | - | | | 7.7 | 313 |
| Infected - no ART | | - | | | | - | | | 6.6 | 170 |
| Uninfected | | - | | | | - | | | 11.8 | 3,954 |
| *Birth/pregnancy history* | | | | | | | | | | |
| Nulli-parous | 1.19 | 0.17-8.16 | 0.19 | 1,046 | 7.76 | 1.72-35.0 | 1.1 | 969 | 0.14 | 1,384 |
| Parous | 1.10 | 0.95-1.26 | 14.7 | 3,114 | 0.96 | 0.82-1.11 | 12.9 | 2,857 | 14.8 | 3,268 |

[1]A recent birth was taken to be one that occurred in the year before interview.

AOR: Odds ratios from multivariable logistic regression adjusted for age-group and site type.

**Table S2** Adjusted odds ratios for sexual abstinence and consistent use of family planning methods in sexually-experienced women aged 15-49 years by marital status: late- and mid- Covid-19 reports compared to before Covid-19, Manicaland, Zimbabwe

| | Sexual abstinence[1] | | | | | | | | | | Consistent use of family planning methods[2] | | | | | | | | | |
|---|---|---|---|---|---|---|---|---|---|---|---|---|---|---|---|---|---|---|---|---|
| | Late-Covid-19 *vs.* Pre-Covid-19 | | | | Mid-Covid-19 *vs* . Pre-Covid-19 | | | | Pre- Covid-19 | | Late-Covid-19 *vs.* Pre-Covid-19 | | | | Mid-Covid-19 *vs* . Pre-Covid-19 | | | | Pre- Covid-19 | |
| Characteristic | aOR | *95% per cent CI* | % | *N* | aOR | *95% per cent CI* | % | *N* | % | *N* | aOR | *95% per cent CI* | % | *N* | aOR | *95% per cent CI* | % | *N* | % | *N* |
| *Sexually-experienced women* | | | | | | | | | | | | | | | | | | | | |
| All | 1.31 | 1.17-1.47 | 20.4 | 3381 | 1.27 | 1.14-1.42 | 19.9 | 3102 | 16.6 | 3679 | 1.05 | 0.96-1.16 | 63.9 | 3381 | 0.82 | 0.74-0.90 | 58.8 | 3102 | 62.1 | 3679 |
| Single | 1.66 | 1.13-2.44 | 54.2 | 192 | 1.77 | 1.19-2.64 | 56.7 | 157 | 44.1 | 295 | 0.94 | 0.62-1.42 | 33.3 | 192 | 0.80 | 0.52-1.24 | 30.6 | 157 | 36.8 | 295 |
| Married | 1.51 | 1.22-1.87 | 7.7 | 2562 | 1.42 | 1.14-1.77 | 7.1 | 2388 | 5.2 | 2803 | 0.97 | 0.87-1.09 | 69.9 | 2562 | 0.74 | 0.66-0.83 | 64.6 | 2388 | 69.2 | 2803 |
| Divorced or separated | 1.31 | 1.01-1.69 | 60.5 | 479 | 1.35 | 1.03-1.76 | 61.1 | 409 | 54.9 | 428 | 1.21 | 0.94-1.57 | 52.6 | 479 | 0.96 | 0.73-1.25 | 46.7 | 409 | 47.5 | 428 |
| Widowed | 1.32 | 0.81-2.14 | 67.6 | 148 | 1.60 | 0.99-2.58 | 73.0 | 148 | 63.4 | 153 | 2.08 | 1.23-3.52 | 37.2 | 148 | 1.46 | 0.90-2.37 | 28.4 | 148 | 22.0 | 153 |
| *Married women* | | | | | | | | | | | | | | | | | | | | |
| Age-group | | | | | | | | | | | | | | | | | | | | |
| 15-19 | 0.62 | 0.28-1.34 | 7.0 | 128 | 1.35 | 0.73-2.50 | 15.2 | 112 | 12.1 | 273 | 0.72 | 0.46-1.11 | 37.5 | 128 | 0.71 | 0.45-1.12 | 36.6 | 112 | 44.7 | 273 |
| 20-24 | 1.27 | 0.75-2.13 | 6.8 | 400 | 2.09 | 1.29-3.39 | 9.9 | 354 | 5.3 | 685 | 0.73 | 0.56-0.93 | 58.0 | 400 | 0.57 | 0.44-0.74 | 52.5 | 354 | 66.0 | 685 |
| 25-34 | 2.59 | 1.63-4.11 | 7.0 | 829 | 1.63 | 0.99-2.66 | 4.6 | 828 | 2.9 | 908 | 0.96 | 0.78-1.19 | 73.7 | 829 | 0.65 | 0.53-0.80 | 65.8 | 828 | 74.6 | 908 |
| 35-49 | 1.46 | 1.06-2.02 | 8.5 | 1205 | 1.23 | 0.88-1.70 | 7.2 | 1094 | 6.0 | 937 | 1.26 | 1.05-1.51 | 74.7 | 1205 | 1.00 | 0.83-1.20 | 70.4 | 1094 | 70.3 | 937 |
| Location | | | | | | | | | | | | | | | | | | | | |
| Urban | 1.55 | 1.02-2.37 | 9.1 | 617 | 1.53 | 0.98-2.38 | 9.4 | 416 | 7.6 | 526 | 1.11 | 0.87-1.42 | 67.1 | 617 | 0.87 | 0.67-1.12 | 60.8 | 416 | 62.0 | 526 |
| Periurban | 2.14 | 1.36-3.37 | 7.6 | 710 | 1.69 | 1.06-2.69 | 6.0 | 745 | 3.7 | 833 | 0.94 | 0.76-1.16 | 69.7 | 710 | 0.85 | 0.68-1.05 | 67.5 | 745 | 69.7 | 833 |
| Agricultural estate | 1.92 | 1.18-3.15 | 7.4 | 605 | 1.48 | 0.88-2.49 | 5.8 | 552 | 3.8 | 625 | 0.91 | 0.71-1.16 | 70.7 | 605 | 0.66 | 0.52-0.85 | 64.7 | 552 | 71.0 | 625 |
| Rural | 0.99 | 0.67-1.47 | 6.7 | 630 | 1.21 | 0.83-1.76 | 7.9 | 675 | 6.8 | 819 | 0.96 | 0.77-1.20 | 72.1 | 630 | 0.63 | 0.50-0.78 | 63.6 | 675 | 71.9 | 819 |

[1]Women reporting not having had sex for 3 months or more were taken to have been abstaining from sexual activity.

[2]Includes modern and traditional methods. Women who reported using a method of contraception most or all of the time were taken to have been using family planning methods consistently.

aOR: Odds ratios from multivariable logistic regression adjusted for age-group and community location.
